# What is the Status of Elderly Abuse in India: A Systematic Review Protocol

**DOI:** 10.1101/2020.11.12.20230284

**Authors:** Raunaq Singh Nagi, Sembagamuthu Sembiah, Anirban Chatterjee, Rudresh Negi

## Abstract

Elder abuse has been identified as one of the major problems of public health globally. Many types of mistreatment practices contribute to elder abuse. Also, the event has been known to occur in community as well as institutional settings such as hospitals and nursing homes. Elder abuse has been shown to affect the quality of life of individuals and increase premature mortality of the elderly. However, majority of elder abuse remains undocumented due to reasons such as fear and lack of awareness of one’s rights among others. Furthermore, the social construct of Indian societies makes the identification and acknowledgement of elder abuse challenging. Although, there have been studies and reports regarding the prevalence of elder abuse in India from various geographical regions, the strength of this evidence is unknown and a general view of prevalence of elder abuse in India is missing.

We propose to undertake a systematic review and metanalysis of the published literature from India regarding the prevalence of elder abuse in any setting. For the purpose we will scan various indexing databases for identification of relevant studies and follow standard protocols and guidelines for formulation of evidence. The results of this study will give an entrusted overview of the status of elder abuse in India and aid policy-making initiatives and interventions to be directed accordingly.

## Introduction

Elder abuse has been recognised as one of the major problems of global public health and a violation of human rights. World Health Organization defines elder abuse as a single or repeated act, or lack of appropriate action, occurring within any relationship where there is an expectation of trust, which causes harm or distress to an older person. (1) Five different types of elder abuse have been identified and addressed in the literature, namely, physical abuse, psychological or verbal abuse, financial abuse, sexual abuse and neglect. (2) The prevalence of elder abuse tends to remain underreported to an extent that only around 4% cases of elder abuse are reported. This is attributed to fear of the elderly while reporting such abuse or unawareness regarding their legal rights and proceedings. (1,3)

Elder abuse has been known to occur in community as well as institutional settings, which include old-age homes, hospitals and nursing homes. Studies have demonstrated a prevalence of 15.7% of self-reported elder abuse in community settings and a staggering prevalence of 64.2% of staff-reported elder abuse in institutional settings. (4,5) Consequences of elder abuse are varied, from increasing morbidity and decreasing the quality of life to reduced survival rates. One study revealed that the rate of premature mortality is double in elderly individuals who suffer abuse. (6) Another study has reported a variety of physical, mental and emotional outcomes that manifest in elders who have experienced abuse. (7)

Research on the topic of elder abuse in India are limited. The issue of elderly abuse in India is slightly unique due to the sandwich structure of the families, where adults take care of the children and older individuals of the family, and the upheld traditional family value system. Abuse remains hidden due to family circumstances, sensitivity of the issue and due to reluctance of victims towards addressing the issue. (8) Several studies have been conducted in different parts of the country in different settings ascertaining the prevalence of elder abuse or its association with physical and psychological manifestations. (3,9,10) A systematic body of evidence regarding the overall prevalence of elder abuse in India is missing.

We propose to undertake synthesis of evidence in the form of a systematic review regarding the prevalence of elder abuse in India. Such evidence would uncover the extent of the problem and attract channelling of resources towards addressing this global public health problem by allowing policy making and targeted interventions.

## Methods

### Protocol and registration

The protocol has been written in accordance to the Preferred Reporting Items for Systematic Reviews and Meta-analysis for Protocols (PRISMA-P) guidelines. (11) The process of screening and selection of studies to be included in the article will be outlined in the form of PRISMA-P flow diagram and PRISMA 2009 statement will be used to formulate the results.

The protocol has been registered on Open Science Framework (OSF) (https://osf.io/t64xq/).

### Search strategy and eligibility criteria

We will use the following freely accessible databases for conducting the search: MEDLINE, JSTOR, ScienceDirect and DOAJ. Reviewer (RSN and SS) conducted a preliminary search using keywords such as “elder abuse”, “elder mistreatment” and other keywords identified through initial screening of retrieved articles. We will use appropriate tools such as Boolean and other operators, medical subheadings and relevant filters to arrive at the final comprehensive search strategy. The search strategy for searching MEDLINE database and corresponding results retrieved on 22.09.2020 are as follows:

1. All field vocabulary: Strategy: (((((elder abuse) OR (elder mistreatment)) OR (elder neglect)) OR (elder maltreatment)) AND (india)) AND (prevalence OR incidence OR epidemiology) # results: 25
2. Controlled vocabulary: Strategy: (“Elder Abuse/epidemiology”[Mesh] OR “Elder Abuse/statistics and numerical data”[Mesh]) AND india # results: 13

We will include all the primary data studies in the review. Review articles will also be included in the initial stages for citation tracking and identification of additional primary studies for inclusion during data extraction and analysis stage. We will include all the studies published since the commencement of the database. Studies published only in English language will be included in the review.

### Training of reviewers

The reviewers to be involved in the screening process of the studies will be versed with the eligibility criteria for inclusion and exclusion of the articles. They will also conduct a practice screening exercise on 50 articles where they analyse the eligibility of these articles for the study. The reviewers will also acquaint themselves with the requirements and usage of software which will be used to conduct the review namely, Mendeley/Zotero and Rayyan and Revman (version 5.4.1, Cochrane).

### Review process

The studies retrieved will be exported in the citation management software Mendeley/Zotero, for organisation of the studies and removal of duplicates. After this stage the studies will be entered in Rayyan for further processes. Two reviewers will individually screen the studies. Firstly, the titles of the retrieved studies will be screened, followed by abstracts. Inclusion and exclusion of studies as per the eligibility criteria will be done at both the stages. Disagreement at any of the stages will be resolved by discussion with an independent third reviewer. Cohen’s kappa statistics will be used to assess the inter-rater agreement. Values at or above 0.80 will be considered to be high degree of agreement. Finally, full-length articles of the studies included after abstract screening will be obtained. These full-length articles will be further evaluated according to the eligibility criteria. Furthermore, citation tracking will be used to identify other relevant studies in the reviews and other such articles. These tracked citations will also be evaluated as per the eligibility criteria. Finally, primary data identified through study retrieval and citation tracking will be included in the systematic review. The whole schematic of the process from retrieval of studies to selection of studies for analysis has been demonstrated in a flowchart (Figure 01).

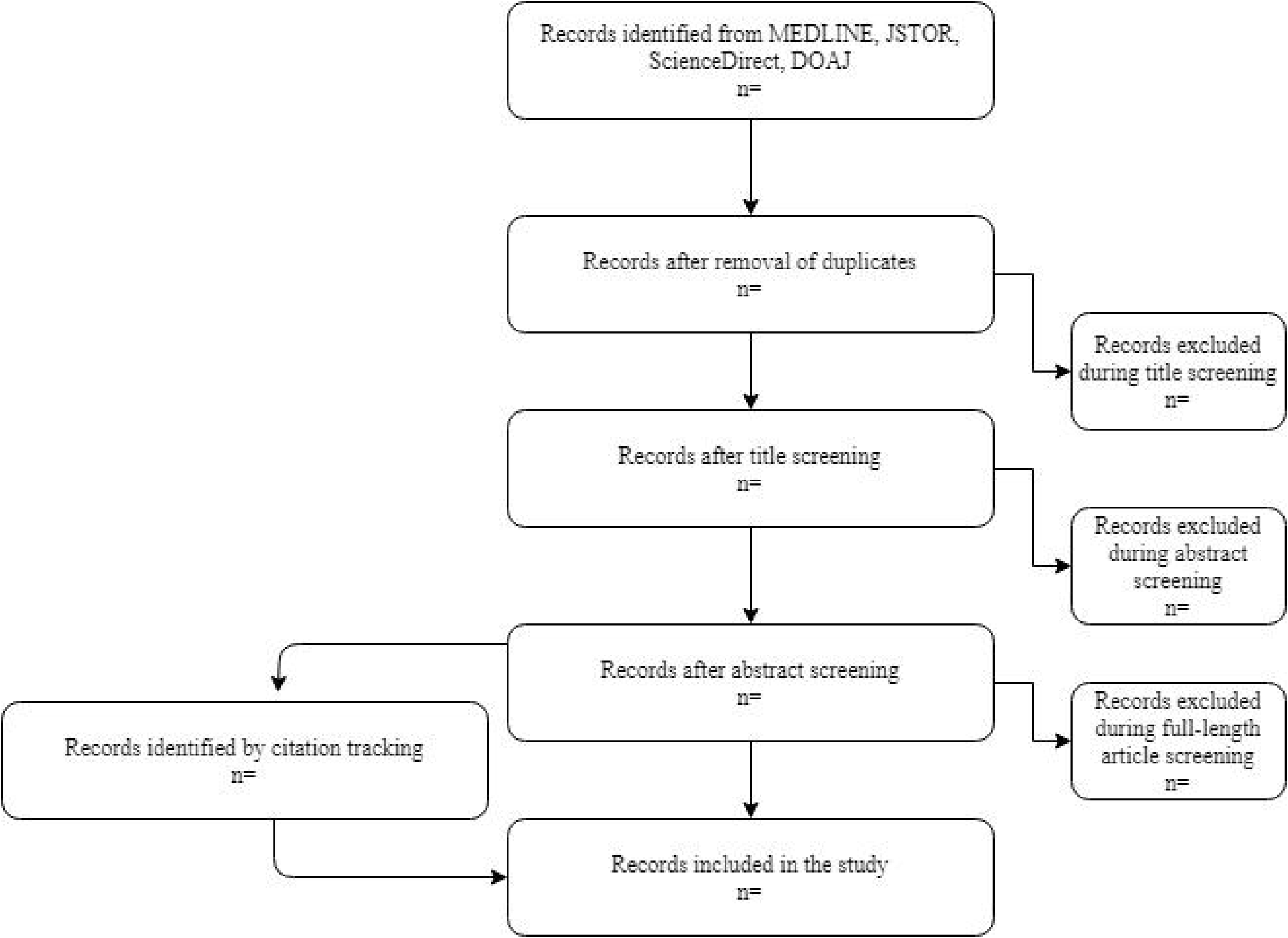

### Data extraction and quality assessment

We will extract the data under the following domains from the full-length studies included in the study, bibliographic data, methodological data and results. Details of author, their country of origin, year of publication, journal details will be included in bibliographic data. Study design, setting, sample size, participant information regarding their age, gender and other characteristics, instruments used for assessment, and its validity, all such details will be included in the methodological data. Results will include information regarding the major findings and any other findings. We will employ Critical Appraisal Checklist for Studies Reporting Prevalence Data by JBI to evaluate the evidence in the included studies. (12)

### Evidence synthesis and statistical analyses

We will briefly report the data descriptively. We also plan to conduct a meta-analysis regarding the overall prevalence of elder abuse in India. We will use Revman for conducting the meta-analysis. Risk of publication bias will be represented in the form of a funnel plot. We will use DerSimonian and Laird inverse variance model for analysis and present the 95% confidence interval (95% CI) of the even rate in the form of a forest plot. We will also assess the variability associated with heterogenicity in the studies included in the analysis using I ^2^ values, where <25% would be considered as low, 25-50% would be considered as medium and >75% would be considered high heterogeneity.

In the event of insufficient number of studies for conducting meta-analysis, we will synthesize the results in the form of forest plots and present a precise and informative description of the analysed studies.

## Data Availability

The data shall be made available up on request made to the authors.

## Financial Statement

This study has not been funded by any agency.

